# Effects of CPAP on OSA-related cardiovascular risk markers: a two-week CPAP withdrawal and re-initiation study

**DOI:** 10.64898/2026.03.10.26348040

**Authors:** Adrien Waeber, Geoffroy Solelhac, Grégory Heiniger, Théo Imler, Monica Betta, Giulio Bernardi, Andrea Faini, Paolo Castiglioni, Carolina Lombardi, Gianfranco Parati, Vincent Pichot, Ali Azarbarzin, Raphael Heinzer

## Abstract

**Background:** The cardiovascular (CV) benefit of CPAP in OSA remains debated and its effects on new OSA-related CV risk markers are unclear. We aimed to quantify short-term CPAP effects on these markers along with vascular and autonomic phenotypes.

**Methods:** In a 2-week withdrawal study, patients on long-standing effective CPAP took part in three visits (V1-V3: on/off/back-on CPAP) with overnight polygraphy followed by vascular and autonomic phenotyping. Co-primary endpoints included endothelial function assessed by flow-mediated dilation (FMD) and baroreflex sensitivity (BRS), hypoxic burden (HB), pulse wave amplitude drop index (PWADi) and spontaneous-PWADi (excl. apnoea-triggered drops), and event-related heart-rate response (ΔHR). Between-visit differences were tested in adjusted mixed models, with visit or within-participant changes in AHI/HB as fixed effects.

**Results:** In 42 participants (61±10 years, 83% male), CPAP withdrawal reinstated OSA (medians [IQR] V1 to V3: AHI 3.9[1.5, 8.8] to 33.4[19.5, 42.1] to 4.0[2.0, 8.8] events/h, HB 4.3[1.1, 8.7] to 51.3[19.7, 83.7] to 2.0[1.2, 6.5] %·min/h, p<0.001) and increased total PWADi (mean±SD 42.25±18.73 to 50.22±17.77 to 41.29±17.14 drops/h, p<0.001), while spontaneous PWADi decreased as respiratory-events recurred (-1.17 drops/h per 10 events/h, p=0.015) along with FMD (3.7±1.9% to 3.2±2.5% to 4.2±2.7%, V2 vs V3 p=0.047). ΔHR and BRS were stable across visits.

**Conclusion:** Short-term CPAP re-initiation improved endothelial function (FMD), with no significant effects on autonomic measures (BRS, ΔHR) or structural vascular indices. This supports a temporal dissociation between rapidly reversible exposure metrics (AHI, HB) and slower dynamics of autonomic markers. Changes in spontaneous PWADi suggests that it may track physiological CPAP benefits beyond indices driven primarily by respiratory-event frequency.

## Introduction

Obstructive sleep apnoea (OSA) is highly prevalent [1, 2] and is associated with increased cardiovascular (CV) morbidity [3–7]. Continuous positive airway pressure (CPAP) is the first-line therapy for moderate-to-severe OSA and improves symptoms and quality of life [8]. However, the extent to which CPAP translates into CV prevention remains controversial, with positive results in prospective clinical cohorts [9, 10] but mixed or neutral effects on CV outcomes have been reported in the main randomized clinical trials [11–13]. These inconsistent findings may be due not only to suboptimal CPAP adherence in these trials but also to limitations of the apnoea-hypopnoea index (AHI) used to select the patients, which incompletely captures the pathophysiological burden relevant to CV risk [14].

Accordingly, recent work has shifted focus toward alternative OSA severity markers that may more accurately reflect the CV burden of the respiratory disorder [15]. These include hypoxic burden (HB) [16], the event-related heart-rate response (ΔHR) [17], and the pulse wave amplitude drop index (PWADi) [18], a photoplethysmography (PPG)-derived index reflecting vascular reactivity. Importantly, these metrics can be extracted not only from polysomnography or respiratory polygraphy but also from nocturnal oximetry, as the oximeter PPG signal contains the information required for their computation [19, 20]. Although several cohorts have reported associations between these markers and cardiovascular outcomes, their modulation by CPAP therapy and physiological correlates are not fully established.

Mechanistically, intermittent hypoxaemia, sleep fragmentation and intrathoracic pressure swings trigger sympathetic activation and haemodynamic stress that may promote autonomic dysregulation and vascular dysfunction, which are two important pathways linking OSA to CV diseases [21, 22]. Repetitive nocturnal blood pressure surges may impair baroreflex function, resulting in reduced baroreflex sensitivity (BRS) and higher blood pressure (BP) [23, 24]. Moreover, hypoxia-reoxygenation can promote oxidative stress and inflammation, impair nitric oxide bioavailability [25] and contribute to endothelial dysfunction, which can be assessed non-invasively by brachial artery flow-mediated dilation (FMD) [26]. Both autonomic and endothelial alterations have been reported to be at least partly reversible with effective CPAP in previous studies [23, 27–29], supporting their role as mechanistically meaningful, treatment-responsive endpoints.

A practical difficulty in conventional CPAP trials initiated in CPAP-naive patients is poor early adherence to therapy. As a result, an alternative experimental approach has been successfully used in prior studies, consisting of recruiting patients with established and adequate CPAP use and temporarily withdrawing CPAP to unmask the physiological consequences of untreated OSA within the same individuals, followed by reassessment after CPAP re-introduction [30]. This within-subject “CPAP withdrawal model” allows efficient evaluation of short-term, reversible changes while reducing confounding by between-person differences in baseline CV risk.

Therefore, the OSAVE trial (Obstructive Sleep Apnoea - Autonomic and Vascular Exploration) aimed to explore the short-term physiological effects of CPAP withdrawal and re-initiation on mechanistic CV pathways and on nocturnal OSA-related severity markers beyond AHI.

Importantly, OSAVE extends prior withdrawal studies by including a treatment re-initiation visit and by integrating concurrent autonomic and vascular phenotyping. The co-primary objectives were to quantify within-subject changes in endothelial function (FMD) and autonomic regulation (supine BRS), as well as CPAP-related changes in nocturnal OSA-related markers, including PWADi, HB and ΔHR.

Secondary objectives were to explore concomitant changes across a broader panel of vascular and autonomic markers. We hypothesised that CPAP withdrawal would be associated with adverse changes in endothelial and autonomic function and with increases in OSA-related physiological markers, and that these alterations might partially reverse after CPAP re-initiation.

## Methods

### Study design and setting

OSAVE is a single-centre, non-randomised, open-label, within-subject before-after trial using a CPAP withdrawal and re-initiation model. As shown in Figure 1, once included, participants completed three study visits: V1 on usual CPAP, V2 after two weeks of CPAP withdrawal, and V3 after two weeks of CPAP re-initiation. The study was conducted at the Centre d’Investigation et de Recherche sur le Sommeil (CIRS), Lausanne University Hospital (CHUV), Switzerland.

**Figure 1:**
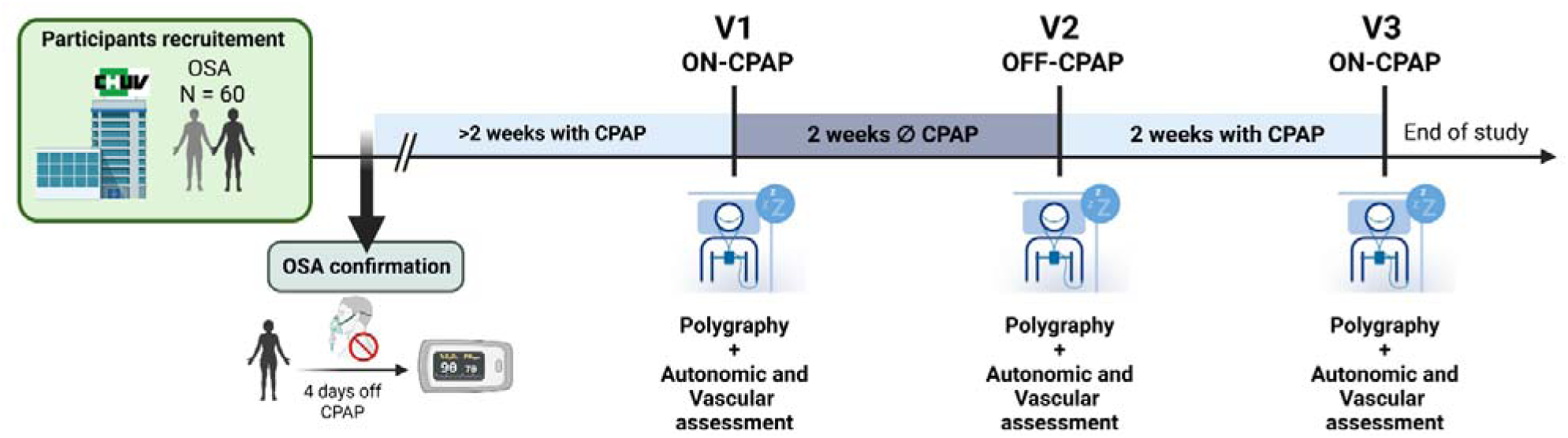
Study design. Following recruitment, participants completed a screening night using an ApneaLink device after 4 days without continuous positive airway pressure (CPAP) to confirm persistent obstructive sleep apnoea off treatment. After screening, participants resumed CPAP for a minimum of 2 weeks before entering the interventional phase. Participants then completed three visits scheduled two weeks apart: visit 1 (V1) on usual CPAP, visit 2 (V2) after 2 weeks of CPAP withdrawal (off CPAP), and visit 3 (V3) after 2 weeks of CPAP re-initiation (on CPAP). At each visit, participants underwent ambulatory overnight respiratory polygraphy followed the next morning by standardised vascular and autonomic assessments, including brachial artery flow-mediated dilation (FMD) and spontaneous baroreflex sensitivity (BRS).

### Participants recruitment and screening

Potential participants were identified from CIRS clinical records based on eligibility criteria and contacted by the study team. Eligible participants were adults (≥18 years) with moderate to severe OSA (AHI ≥ 15/h) and established CPAP therapy for at least six months (defined as device-derived adherence > 70% of nights with CPAP use and mean use > 4h/night over the preceding months).

Key exclusion criteria were professional drivers and conditions or treatments expected to substantially affect endothelial or autonomic function (diabetes, heart failure, chronic kidney disease, morbid obesity ≥40 kg/m², atrial fibrillation, pregnancy/lactation, alpha- or beta-blockers, excessive alcohol consumption (>14 standard units/week), dementia). Written informed consent was obtained prior to study procedures.

As shown in Figure 1, a first screening procedure was performed to confirm persistence of OSA upon CPAP withdrawal (given that diagnosis and CPAP initiation could have occurred years earlier), participants discontinued CPAP for four days and underwent ambulatory screening using ApneaLink™ (ResMed Corp., San Diego, CA, USA). Participants with AHI <15 events/h, were not included in the intervention phase. Baseline demographic and clinical characteristics of participants with persistent OSA were collected at this inclusion visit (see “Study visit procedures”). Then, they resumed CPAP for at least two weeks before entering the intervention phase (i.e., before V1) to ensure a stable treated-state assessment.

### Intervention

The intervention consisted of a two-week CPAP withdrawal period (between V1 and V2), followed by two weeks of usual CPAP use (between V2 and V3). Participants were instructed not to use CPAP during the withdrawal period and to resume their usual CPAP settings thereafter, adherence to CPAP use was documented using device downloads (average nightly use and % nights used)

### Study visits procedures

Each visit (V1, V2 and V3) comprised an ambulatory overnight polygraphy followed the next morning by questionnaires and a standardised panel of vascular and autonomic assessments, as detailed below. Morning assessments were performed exactly two weeks apart and at the same time for each participant, between 08 and 10 am depending on individual wake-up time. Participants fasted for ≥6 h, avoided strenuous exercise for ≥24 h, and refrained from alcohol/caffeine for ≥12 h and smoking for ≥6 h prior to testing.

### Endpoints

<u>Co-primary endpoints</u> were (1) endothelial function assessed by FMD, (2) autonomic regulation assessed by supine BRS and (3) nocturnal OSA-related markers (PWADi, HB and ΔHR). <u>Secondary endpoints</u> included peripheral arterial stiffness and bilateral common carotid artery assessment, seated BRS and additional baroreflex indices (Baroreflex effectiveness index (BEI), positive and negative latencies), nocturnal and diurnal heart rate variability (HRV), cold pressor test (CPT) responses, and Epworth Sleepiness Scale (ESS).

### Sleep recordings and OSA-related markers

Ambulatory nocturnal polygraphy was performed using a NOX T3 recorder (Nox Medical, Reykjavik, Iceland), including nasal pressure (airflow), thoracoabdominal effort belts, one lead ECG, and pulse oximetry using a Nonin WristOx2 3150 (Nonin, Plymouth, USA). Pulse oximetry specifications are described in the online supplement.

Respiratory events were scored according to the 2023 AASM (recommended) criteria [31] using Noxturnal Sleep Study Software (Nox Medical, Reykjavik, Iceland).

OSA-related CV risk markers were extracted from the same recordings. The PWADi was computed automatically using a validated MATLAB algorithm developed in our laboratory [32], detecting >30% drops in PPG pulse wave amplitude relative to a dynamically updated baseline. PWADi was defined as the number of detected drops per hour of recording. “Spontaneous” PWADi was defined as the frequency of pulse wave amplitude drops not preceded by a scored respiratory event within the preceding 10 s, expressed per hour of analysed recording time after excluding all 10 sec windows associated with respiratory events. HB and ΔHR were computed using MATLAB implementations adapted from published methods [16, 17]. Briefly, HB was calculated as the area between an adaptive expected baseline saturation and the observed post-event desaturation curve for each respiratory event, summed across the night and expressed per hour of recording (%·min/h). ΔHR quantified the event-related pulse-rate acceleration as the difference between the minimum heart rate during the respiratory event and the maximum heart rate within adaptative post-event search window (beats/min), derived from the PPG signal. ΔHR was computed only for recordings containing >50 respiratory events, consistent with the approach used by Azarbarzin et al. [17].

### Vascular assessments

#### Flow-mediated dilation

Brachial artery FMD was assessed using high-resolution duplex ultrasound (Philips InnoSight, linear probe L12-4) performed using edge-detection software (FMD Studio©, Quipu, Pisa, Italy) with a stereotactic probe holder to reduce operator dependence, following expert recommendations [26]. Briefly, baseline brachial artery diameter was recorded for 1 min prior to cuff inflation. A distal forearm cuff was inflated to 50 mmHg above systolic BP for 5 min and then rapidly deflated, arterial diameter was continuously tracked for 4 min post-deflation to determine peak diameter and time-to-peak. Detailed methods, including quality control and missing data (Table S1) are described in the online supplement.

#### Peripheral arterial stiffness

Peripheral arterial stiffness was assessed using the pOpmètre® system (pOpscope software v3.1.67, Axelife, Paris, France). Pulse wave velocity was derived from simultaneous PPG recordings obtained at the finger and at the toe on the right side over a short recording period. The device computes the finger-toe pulse transit time from the foot of the PPG waveforms and estimates finger-toe pulse wave velocity as travelled distance divided by transit time, with travelled distance estimated from participant height.

Using the same software, central systolic and diastolic blood pressures were estimated from the recorded signals using the manufacturer’s proprietary algorithm [33], and these values were subsequently used for carotid distensibility calculations (see below).

#### Bilateral carotid stiffness

Bilateral carotid stiffness was assessed by ultrasound (Philips InnoSight, linear probe L12-4) using Carotid Studio© (Quipu, Pisa, Italy), enabling continuous tracking of common carotid artery (CCA) diameter and intima-media thickness (IMT, mm). Estimated central systolic and diastolic blood pressures were used for pressure-calibrated stiffness indices including local pulse wave velocity (m/s), compliance, distensibility and Young’s modulus. Detailed methods are included in the online supplement.

### Autonomic assessments

#### Baroreflex sensitivity (BRS)

Beat-to-beat finger arterial pressure was recorded using a volume-clamp device (Finapres NOVA, Finapres Medical Systems, The Netherlands) in supine and seated positions (10 min each), synchronised with a 5-electrodes ECG. The sensitivity of the baroreflex control of heart rate was estimated by the transfer function technique (HLF) [34, 35]. BRS calculation is further described in the online supplement. BEI (which provides information on how often the baroreflex takes control of the sinus node in response to spontaneous BP changes) and baroreflex latencies were derived using the sequence technique [36, 37]. Diurnal ECG-derived HRV indices were computed from the same recordings.

#### Cold pressor test (CPT)

The CPT consisted of immersing the right hand in iced water (0-1°C) for 120 s, up to the wrist following at least 5 min of supine baseline recording. Heart rate and beat-to-beat blood pressure were continuously recorded on the left hand using the Finapres NOVA, and digital PPG was recorded using a Nonin WristOx2 3150 pulse oximeter. Signals were acquired throughout baseline, cold exposure, and a 5-minute recovery period. CPT responses were summarised as peak changes from baseline (HR, Systolic BP and Diastolic BP).

#### Heart rate variability (HRV)

Nocturnal ECG-derived RR intervals were analysed using HRVanalysis (HRV Analysis v1.2, ANSLabTools, Université Jean Monnet Saint-Étienne, France). RR series underwent visual inspection and artefact handling using the software’s built-in procedures, including automated detection and correction of ectopic or erroneous beats. Standard HRV indices were then derived from the cleaned RR series, including time-domain (SDNN, RMSSD) and frequency-domain (VLF, LF, HF, LF/HF) measures, as well as nonlinear indices (acceleration and deceleration capacity, SampEn and DFA α1 and α2). HRV analyses were restricted to the nocturnal period (night-time window entered for each recording).

### Baseline characteristics

Baseline demographic and clinical characteristics were collected once at inclusion into the interventional phase (i.e., after return of the ApneaLink™ device and confirmation of persistent OSA). Data included medical history and CV risk factors, smoking status and alcohol intake, symptom questionnaires (Epworth Sleepiness Scale and Insomnia Severity Index [38, 39]), and a standardised clinical examination with anthropometry, office blood pressure and Mallampati score [40]. CPAP treatment history and device-derived adherence metrics over the preceding 6 months (average nightly use, percentage of nights used, and residual AHI on CPAP) were extracted. A fasting blood sample was obtained at V1 to assess cardiometabolic biomarkers (lipid profile, fasting glucose and HbA1c), inflammatory status (C-reactive protein) and renal-related markers (creatinine, uric acid).

### Sample size

The primary aim of this mechanistic study was to characterise within-subject changes in endothelial function (FMD) and autonomic regulation (BRS) across CPAP withdrawal and re-initiation, and to explore their associations with longitudinal changes in OSA-related exposure metrics. The sample size (65 planned completers) was determined based on feasibility considerations and on the ability to detect moderate-to-large within-subject effects in mixed models. Given the multidimensional assessment and final sample size, analyses should be interpreted as hypothesis-generating. Due to recruitment constraints within the planned timeframe and available resources, 60 patients were recruited, and 42 participants completed all three visits (see Figure 2). Analyses are considered exploratory and missingness is reported for each endpoint (Table S1).

**Figure 2:**
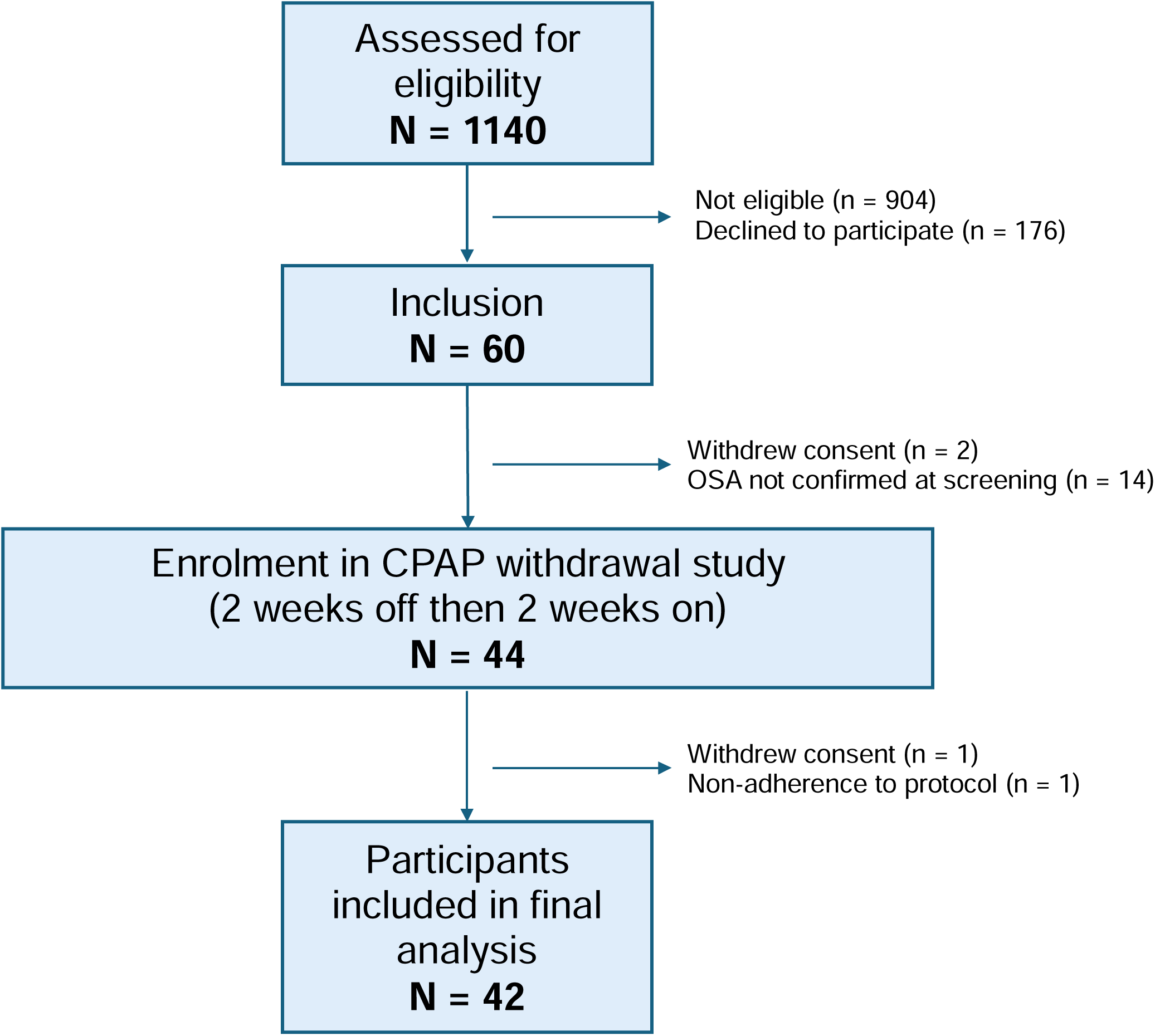
Flow chart. Patients followed at the Centre d’Investigation et de Recherche sur le Sommeil, for CPAP therapy were assessed for eligibility and included between 12 January 2024 and 12 May 2025. Included participants then underwent an ambulatory CPAP discontinuation screening period to confirm persistent obstructive sleep apnoea (OSA) off treatment. Participants with an apnoea-hypopnoea index (AHI) <15 events/h on ApneaLink™ after 4 nights without CPAP were excluded. Participants who met the screening criterion were subsequently enrolled into the interventional phase. The final analysis included those who completed all three visits with adherence to the CPAP withdrawal and re-initiation protocol.

### Statistical analysis

Continuous variables are reported as mean ± SD or median [IQR], depending on distribution, and categorical variables as n (%). Within-subject changes across visits (V1, V2 and V3) were assessed using linear mixed-effects models adjusted for age, sex and BMI, with a participant-level random intercept and prespecified pairwise contrasts (V1 vs V2, V2 vs V3, and V1 vs V3). To obtain a more granular estimate of treatment “effectiveness” and to consider variability in AHI reduction beyond the categorical visit factor, we additionally modelled physiological outcomes as a function of longitudinal OSA severity using a within-between decomposition of AHI and HB. Specifically, AHI and HB were decomposed into a within-person component (deviation from each participant’s mean across visits) and a between-person component (participant-specific mean), both entered simultaneously in the mixed model. The coefficient of primary interest was the within-person effect, quantifying how changes in AHI or HB within the same individual were associated with concurrent changes in vascular and autonomic parameters, while accounting for between-person differences. Co-primary endpoints were tested at a two-sided α=0.05 each without multiplicity correction, whereas secondary outcomes were considered exploratory and were adjusted using a false discovery rate procedure (Benjamini-Hochberg) applied within predefined outcome domains (autonomic vs vascular). Missing data were not imputed.

### Software

Sleep-marker extraction was performed in MATLAB (R2023a). Statistical analyses were performed in R, version 4.4.2.

### Ethics, monitoring and safety

The study was conducted in accordance with the Declaration of Helsinki, ICH-GCP and Swiss regulations (ClinO). The protocol was approved by the local ethics committee (CER-VD, project ID 2023-01043) and the trial was registered at ClinicalTrials.gov (NCT05920083). External monitoring was performed independent from the investigative team, according to an a priori monitoring plan, including verification of informed consent, eligibility, protocol compliance, adverse event reporting and source data verification for prespecified critical variables. Adverse events were collected during the CPAP withdrawal period, with moderate-to-severe events assessed for relatedness to the intervention, serious adverse events were monitored throughout the study and reported according to Swiss requirements.

## Results

### Participants and protocol adherence

Of 60 participants recruited, 42 completed all three visits and were included in the main analyses (Figure 2). Baseline characteristics are summarised in Table 1. Participants were predominantly male (35/42, 83%) with a mean age of 61 ± 10 years. They had a high cardiometabolic risk burden including obesity (mean body mass index 30.3 ± 4.7 kg/m²), known hypertension (22/42, 52%), known dyslipidaemia (15/41, 37%), and 26/42 (62%) were current or former smokers. Seven participants had prior CV events (ischemic heart disease, stroke or peripheral arterial disease).

**Table 1:** Baseline characteristics of the OSAVE cohort. Baseline characteristics were collected at inclusion into the interventional phase (after confirmation of persistent OSA off CPAP) and blood sample was collected at V1 as specified in Methods. ^1^Values are mean ± SD, median [IQR], or n (%), as appropriate.

| Group | Characteristic | N = 42<sup>1</sup> | Missing, n |
| --- | --- | --- | --- |
| **Demographics & comorbidities** | Age, years | 61 ± 10 | 0 |
|  | Women | 7 (17%) | 0 |
|  | Previous CV disease | 7 (17%) | 0 |
|  | Known hypertension | 22 (52%) | 0 |
|  | *Antihypertensive treatment* | 19 (45%) | 0 |
|  | Known dyslipidaemia | 15 (37%) | 1 |
|  | *Lipid-lowering treatment* | 9 (21%) | 0 |
|  | Smoker or ex-smoker | 26 (62%) | 0 |
|  | Alcohol, units per week | 5.4 ± 4.7 | 0 |
|  | Epworth Sleepiness Scale | 7.8 ± 3.9 | 1 |
|  | Insomnia Severity Index | 7.4 ± 5.4 | 13 |
| **Anthropometry & blood pressure** | Body mass index, kg/m<sup>2</sup> | 30.3 ± 4.7 | 0 |
|  | Waist circumference, cm | 106 ± 12 | 0 |
|  | Hip circumference, cm | 106 ± 9 | 0 |
|  | Neck circumference, cm | 39.27 ± 3.12 | 0 |
|  | Office systolic BP, mmHg | 135 ± 14 | 0 |
|  | Office diastolic BP, mmHg | 86 ± 11 | 0 |
|  | Resting heart rate, bpm | 72 ± 13 | 0 |
| **OSA diagnosis & CPAP treatment** | AHI at diagnostic, events/hour | 42 ± 21 | 0 |
|  | Total time on CPAP, months | 91 ± 59 | 0 |
|  | 6-month mean CPAP use, hours/night | 6.08 ± 1.41 | 0 |
|  | 6-month CPAP nights used, % | 94 [80, 98] | 0 |
|  | 6-month mean residual AHI, events/hour | 1.40 [0.80, 3.00] | 0 |
| **Blood biomarkers** | Total cholesterol, mmol/L | 5.22 ± 0.97 | 0 |
|  | HDL cholesterol, mmol/L | 1.40 ± 0.48 | 0 |
|  | LDL cholesterol, mmol/L | 3.13 ± 0.90 | 0 |
|  | Triglycerides, mmol/L | 1.51 ± 0.70 | 0 |
|  | C-reactive protein, mg/L | 2.29 ± 2.10 | 0 |
|  | Fasting plasma glucose, mmol/L | 5.40 ± 0.67 | 0 |
|  | HbA1c, % | 5.46 ± 0.35 | 0 |
|  | Creatinine, µmol/L | 85 ± 14 | 0 |
|  | Uric acid, µmol/L | 362 ± 59 | 0 |

**Abbreviations:** CV, cardiovascular; BP, blood pressure; OSA, obstructive sleep apnoea; CPAP, continuous positive airway pressure; AHI, apnoea-hypopnoea index; HDL, high-density lipoprotein; LDL, low-density lipoprotein; HbA1c, glycated haemoglobin.

AHI at diagnostic sleep study was (mean±SD) 42 ± 21 events/h. Prior CPAP treatment was long-standing (mean ± SD duration 91 ± 59 months) with a 6-month mean use 6.08 ± 1.41 h/night, % of nights used 94 [80, 98] and a median residual AHI 1.40 [0.80, 3.00] events/h).

During the interventional phase, CPAP utilisation was controlled with objective CPAP usage data at V1 (14 nights mean use 6.56 h/night, 96.3% of nights) and V3 (14 nights mean use 6.7 h/night, 98.4% of nights), and complete 14 nights-withdrawal at V2 (0 h/night, 0% of nights).

### Recurrence and reversal of OSA severity

CPAP withdrawal induced a marked recurrence of OSA and hypoxaemia (Figure 3 and Table 2). Median AHI increased from 3.90 [1.50, 8.80] events/h on CPAP at V1 to 33.35 [19.45, 42.10] events/h off CPAP at V2 (p-value <0.001), and returned to 4.00 [1.95, 8.82] events/h after CPAP re-initiation at V3 (V2 vs V3 p-value <0.001, V1 vs V3 p-value =0.985). Hypoxic burden increased substantially during withdrawal, from 4.33 [1.11, 8.67] %·min/h at V1 to 51.32 [19.70, 83.66] %·min/h at V2 (p-value <0.001), and decreased again at V3 (1.97 [1.21, 6.49] %·min/h, V2 vs V3 p-value <0.001, V1 vs V3 p-value =0.839). Similar patterns were observed for ODI, mean SpOL, T90, and mean desaturation amplitude (all V1 vs V2 and V2 vs V3 p-value <0.001).

**Figure 3:**
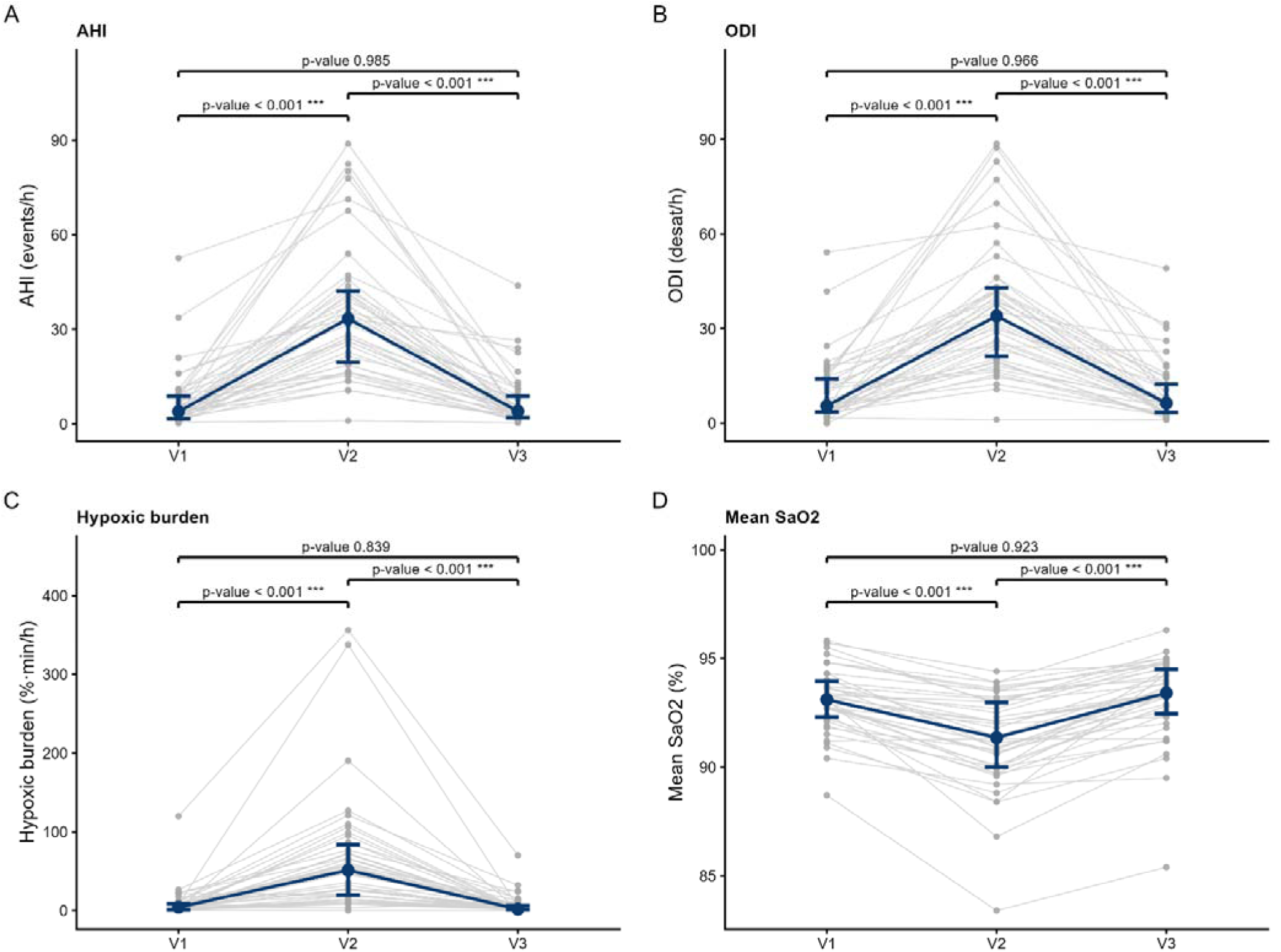
OSA severity across study visits. Key polygraphy outcomes (including **A** apnoea-hypopnoea index, AHI; **B** oxygen desaturation index, ODI; **C** hypoxic burden (HB) and **D** mean oxygen saturation, mean SpO) are presented for V1 (on CPAP), V2 (off CPAP), and V3 (on CPAP). Blue markers represent the median with interquartile range (IQR). Individual participant values are shown as grey dots, connected across visits by grey lines. Pairwise p-values correspond to linear mixed-effects model contrasts adjusted for age, sex, and body mass index (BMI).

**Table 2:**
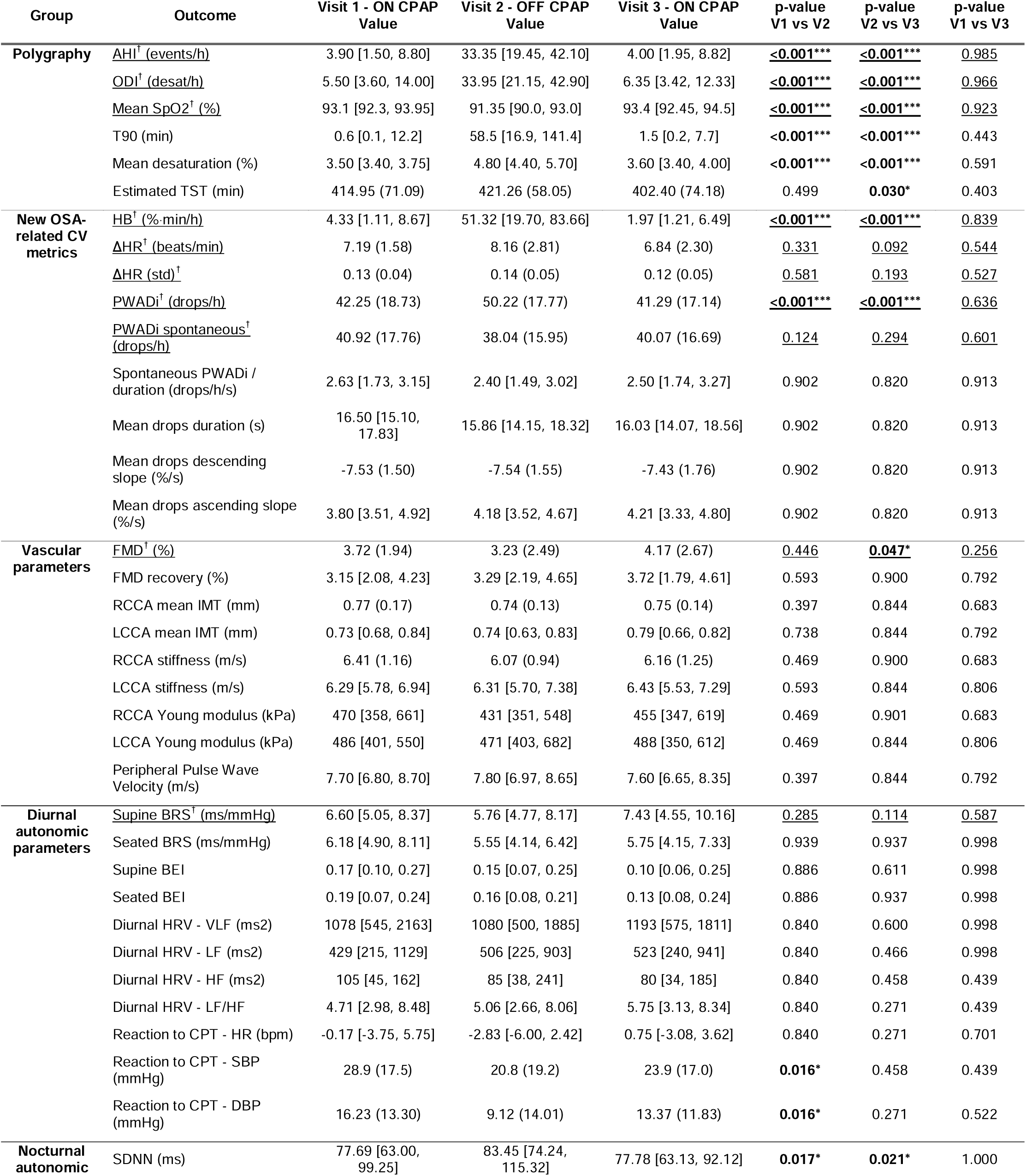
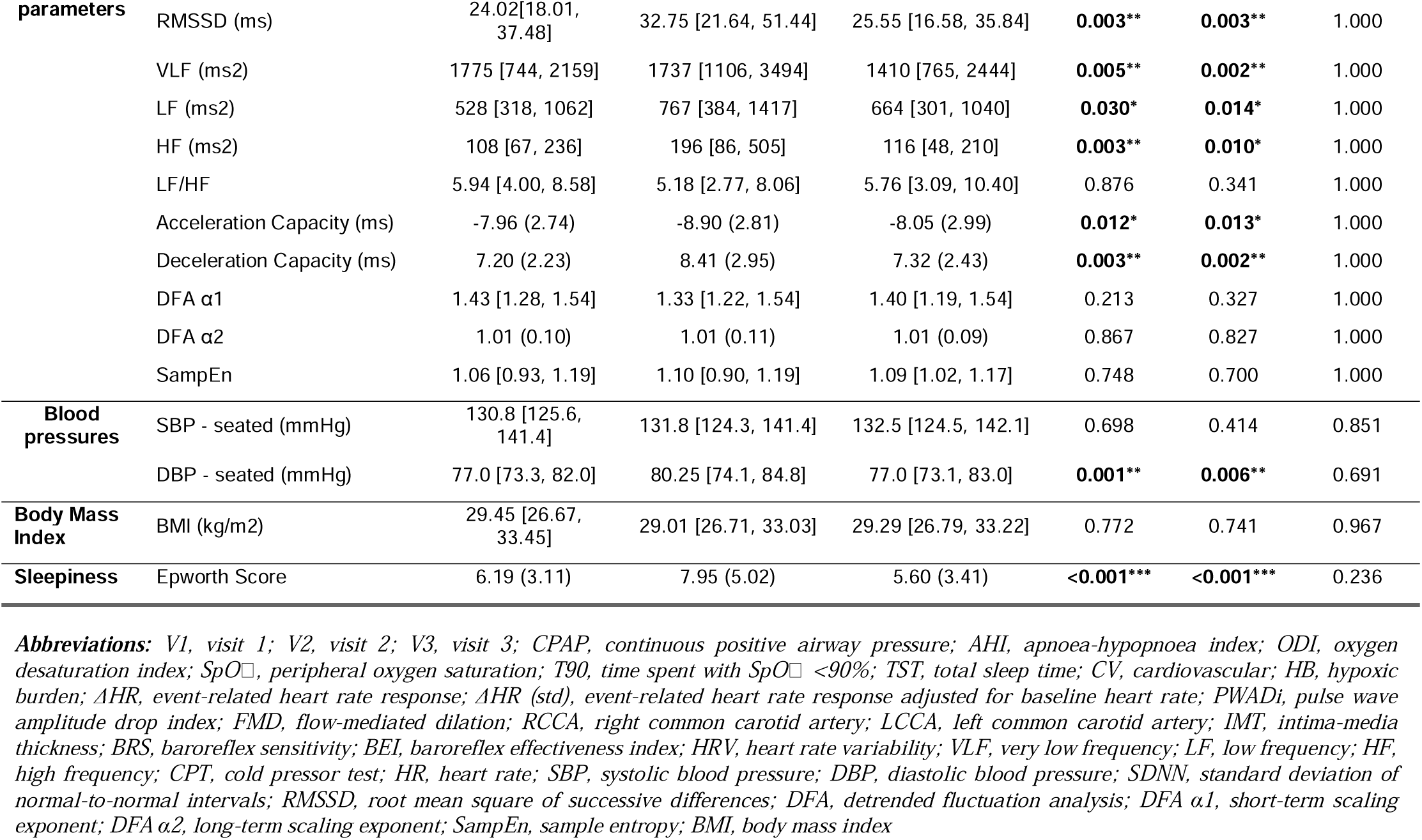
primary and secondary endpoints across study visits. Outcomes marked with † and underlined have unadjusted p-values (primary endpoints); all other outcomes have Benjamini-Hochberg FDR-adjusted p-values within their outcome domain (Group). Descriptive values are mean (SD) or median [IQR] as appropriate. Pairwise between-visit differences were estimated using a linear mixed-effects model adjusted for age, sex and BMI, with a participant-specific random intercept; p-values are derived from these LMM-adjusted contrasts. P-values Significance: * p<0.05, ** p<0.01, *** p<0.001.

### Co-primary vascular and autonomic endpoints

As shown in Figure 4, endothelial function, assessed by brachial artery FMD, was non significantly lower at V2 than at V1 (V1: 3.72 ± 1.94%, V2: 3.23 ± 2.49%, V1 vs V2 p-value = 0.446) and increased significantly after CPAP re-initiation (V3: 4.17 ± 2.67%, V2 vs V3 p-value = 0.047)

**Figure 4:**
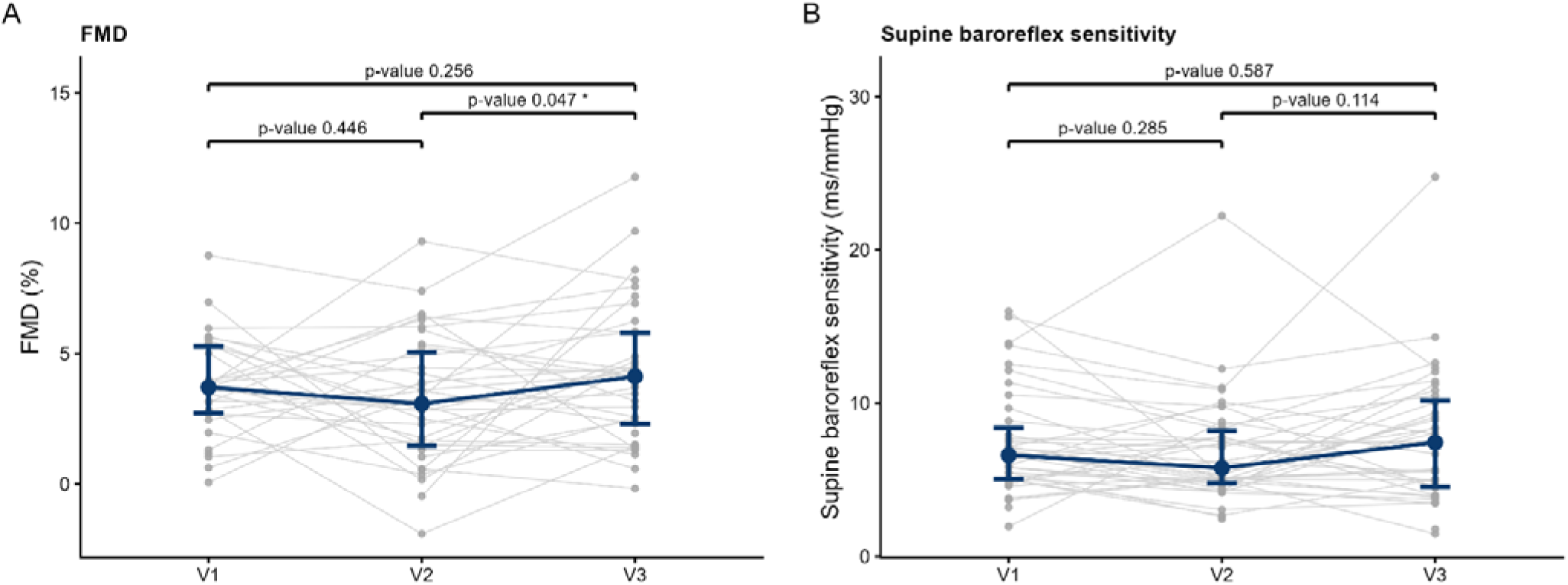
Endothelial function and baroreflex sensitivity across study visits. **A** Brachial artery flow-mediated dilation (FMD, %) and **B** baroreflex sensitivity (BRS, ms/mmHg) are shown at V1 (on CPAP), V2 (off CPAP), and V3 (on CPAP). Blue markers represent the median with interquartile range (IQR). Individual participant values are shown as grey dots, connected across visits by grey lines. Pairwise p-values correspond to linear mixed-effects model contrasts adjusted for age, sex, and body mass index (BMI).

Although not significant, supine baroreflex control of heart rate, BRS, tended to be lower at V2 compared to V1 or V3 (V1: 6.60 [5.05, 8.37] ms/mmHg, V2: 5.76 [4.77, 8.17], V3: 7.43 [4.55, 10.16], all pairwise comparisons p-value ≥0.114).

### New OSA-related CV risk markers

#### PWAD indices

Total PWADi differed across visits (Figure 5 and Table 2). PWADi increased during CPAP withdrawal (V1: 42.25 ± 18.73 drops/h, V2: 50.22 ± 17.77, p-value <0.001) and returned towards baseline after CPAP re-initiation (V3: 41.29 ± 17.14, V1 vs V3 p-value =0.636). By contrast, spontaneous PWADi showed a non-significant trend in the opposite direction, with a lower mean spontaneous PWADi at V2 compared to on-CPAP visits (V1: 40.92 ± 17.76, V2: 38.04 ± 15.95, V3: 40.07 ± 16.69, all pairwise p-value ≥0.124).

**Figure 5:**
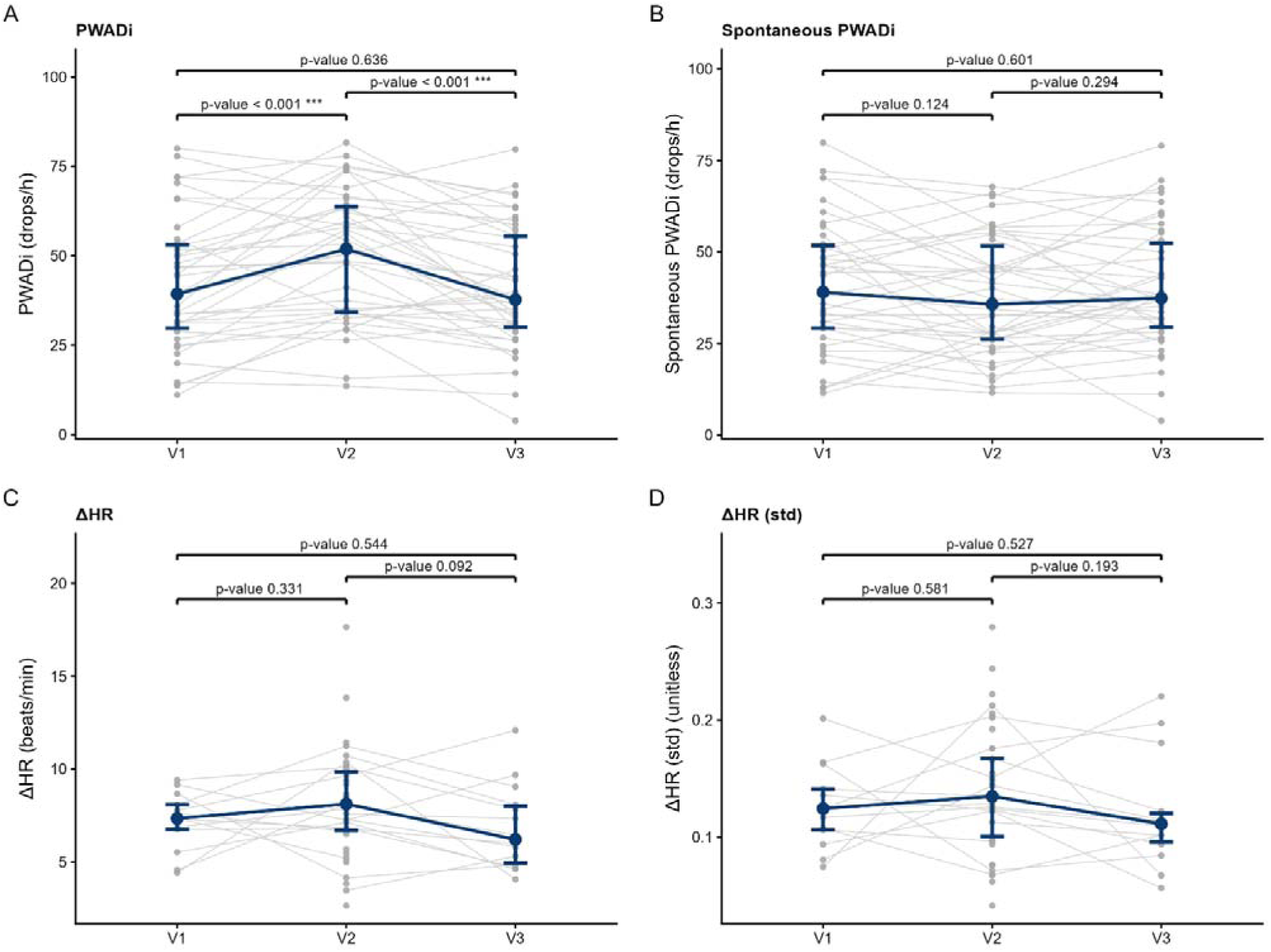
PWADi and ΔHR across study visits. **A** Pulse wave amplitude drop index (PWADi, drops/h), **B** “spontaneous” PWADi, **C** event-related heart rate response (ΔHR, beats/min) and **D** ΔHR adjusted for baseline heart rate are shown for V1 (on CPAP), V2 (off CPAP), and V3 (on CPAP). Blue markers represent the median with interquartile range (IQR). Individual participant values are shown as grey dots, connected across visits by grey lines. Pairwise p-values correspond to linear mixed-effects model contrasts adjusted for age, sex, and body mass index (BMI).

Spontaneous PWADi normalised by drop duration and drop morphology parameters (mean drop duration, descending slope, and ascending slope) also did not differ across visits (all pairwise p-value ≥0.820, Table 2).

### Δ*HR*

Event-related heart rate response did not show clear between-visit differences (ΔHR: 7.19 ± 1.58, 8.16 ± 2.81, and 6.84 ± 2.30 beats/min at V1, V2 and V3, respectively, all p-value ≥0.092, Table 2). ΔHR showed substantial missingness at V1 and V3 because recordings with fewer than 50 respiratory events across the night were excluded from its computation (Table S1).

### Vascular and autonomic secondary endpoints

Structural vascular parameters, including carotid intima-media thickness, carotid stiffness and Young’s modulus, and peripheral pulse wave velocity, did not differ across visits (all pairwise p-value ≥0.397, Table 2).

Baroreflex effectiveness index and latencies remained unchanged across study visits (Table 2 and S2). Nocturnal HRV indices showed heterogeneous changes across visits. SDNN, RMSSD, and frequency-domain components (VLF, LF, HF) increased at V2 compared with V1 and V3 (all p < 0.03). Acceleration and deceleration capacities also improved at V2 (all p < 0.013). By contrast, DFA α1 and α2, sample entropy, and the LF/HF ratio did not differ across visits, and diurnal HRV indices showed no significant changes.

Blood pressure responses to the CPT were significantly attenuated during withdrawal. Peak systolic and diastolic blood pressure responses were lower at V2 than at V1 (ΔSBP: 28.9 ± 17.5 vs 20.8 ± 19.2 mmHg, p-value = 0.016, ΔDBP: 16.23 ± 13.30 vs 9.12 ± 14.01 mmHg, p-value = 0.016), with only partial reversal after CPAP re-initiation.

Finally, seated diastolic BP increased significantly after CPAP withdrawal and decreased after re-initiation (77.00 [73.25, 82.00] mmHg at V1, 80.25 [74.12, 84.75] at V2, 77.00 [73.12, 83.00] at V3, V1 vs V2 p-value =0.001, V2 vs V3 p-value =0.006), whereas seated systolic BP did not differ across visits. Mean blood pressure derived from continuous finger arterial pressure recordings (Finapres) over a 10-min period is detailed in the online supplement.

### Within-person severity-response analyses

In within-between models relating longitudinal outcomes to within-person changes in OSA severity (Table 3 and table S3), increases in AHI were associated with higher total PWADi (+2.70 drops/h per 10 events/h, p-value <0.001) but lower spontaneous PWADi (-1.17 drops/h per 10 events/h, p-value =0.015).

**Table 3:**
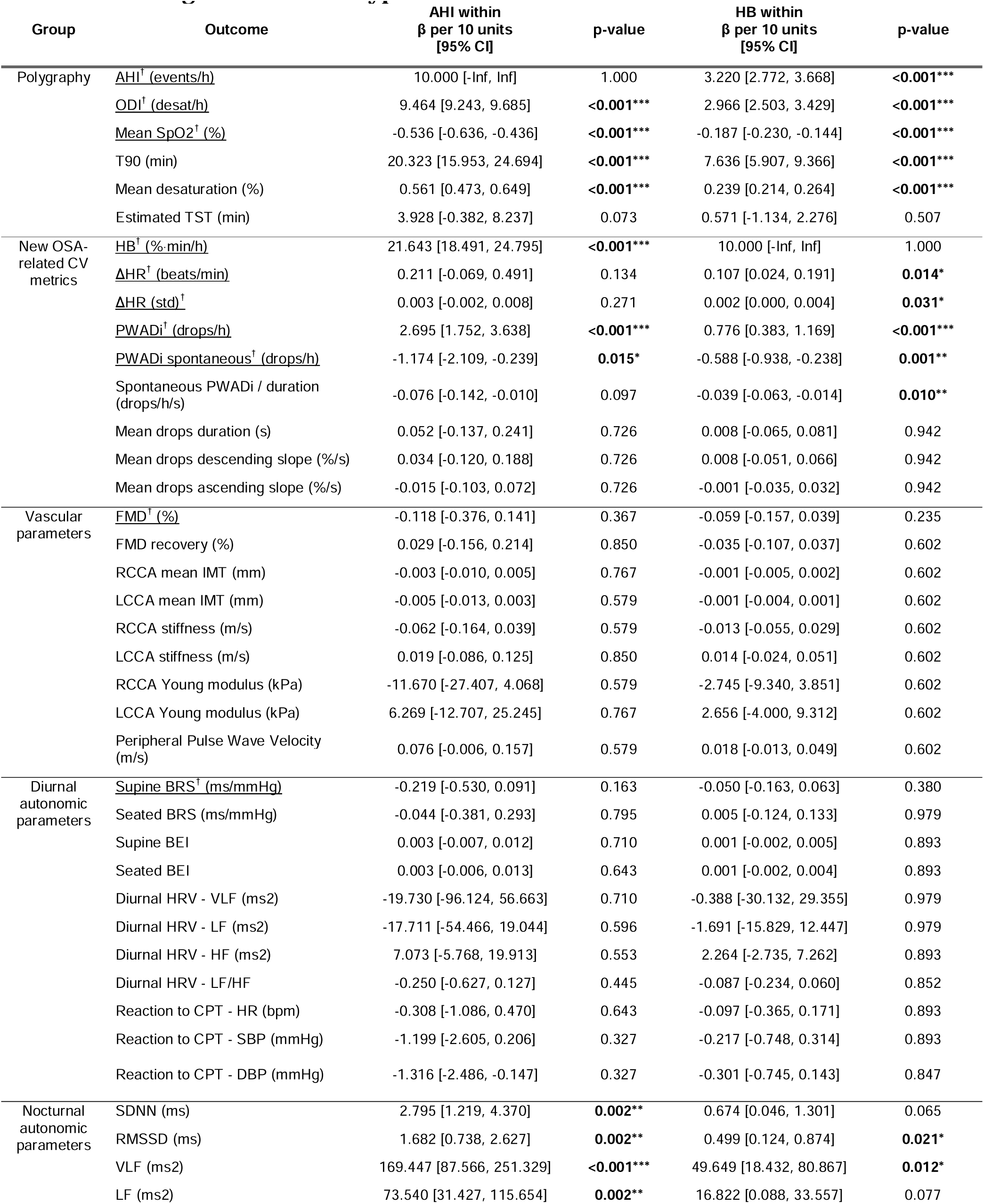
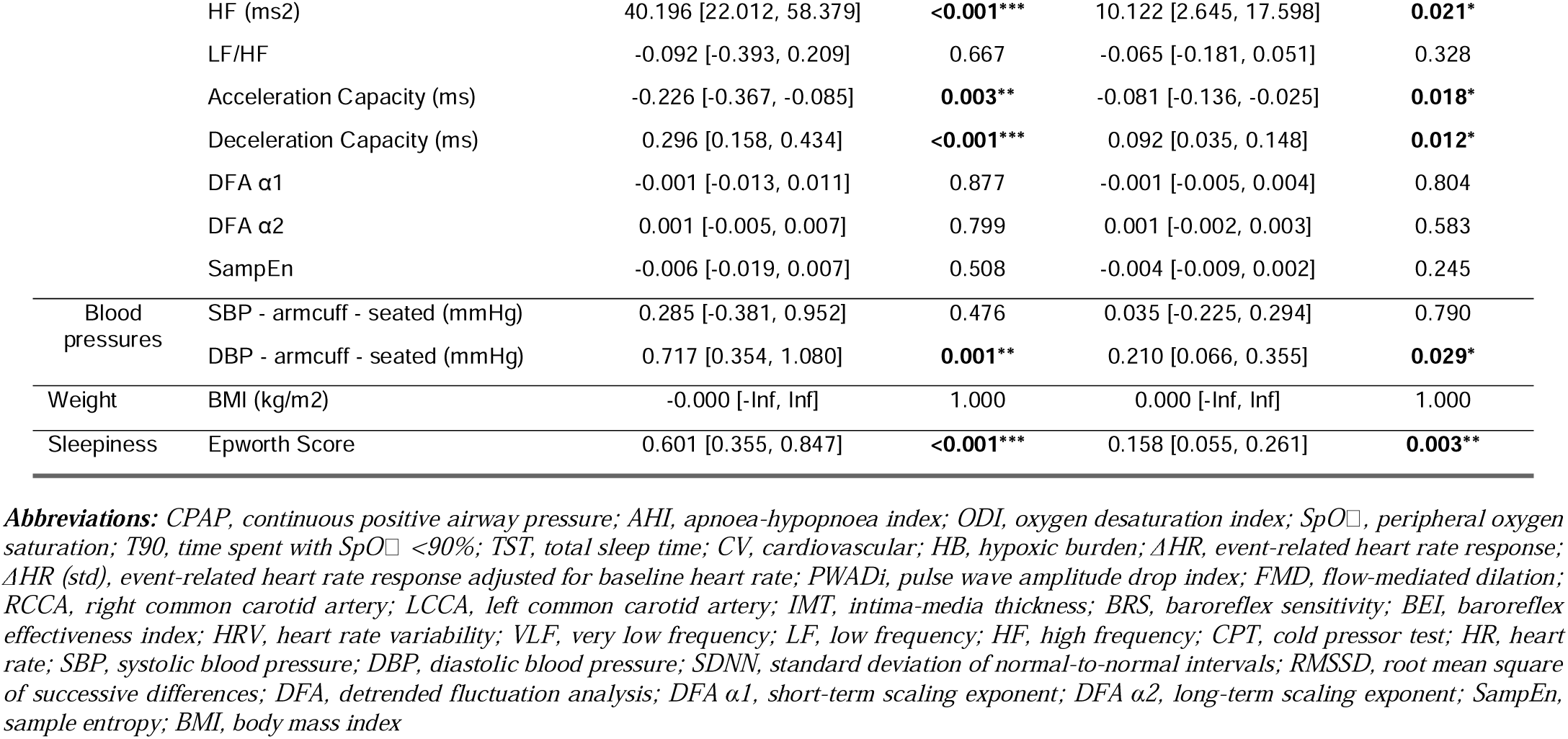
Within-patient associations of primary and secondary endpoints with changes in AHI and hypoxic burden. Associations were estimated using mixed models adjusted for age, sex and BMI, with a participant-specific random intercept, including a within–between decomposition of AHI and HB: the within-person component represents deviation from each participant’s mean across visits, and the between-person component represents participant-specific means. β coefficients are shown per 10-unit increase in within-person AHI (events/h) or within-person HB (%·min/h). Outcomes marked with † and underlined have unadjusted p-values (primary endpoints); all other outcomes have Benjamini-Hochberg FDR-adjusted p-values within their outcome domain (Group). P-values Significance: * p<0.05, ** p<0.01, *** p<0.001.

Within-person increase in hypoxic burden was associated with higher ΔHR (+0.107 beats/min per 10 %·min/h, p-value =0.014) and ΔHR adjusted for baseline heart rate (+0.002 per 10 %·min/h, p-value =0.031), higher total PWADi (+0.776 drops/h per 10 %·min/h, p-value <0.001), and lower spontaneous PWADi (-0.588 drops/h per 10 %·min/h, p-value =0.001) as well as lower spontaneous PWADi normalised by duration (-0.039 drops/h/s per 10 %·min/h, p-value =0.010).

Within-person changes in AHI or HB were not significantly associated with FMD or supine BRS (all p-value > 0.163, Table 3).

### Symptoms and adverse events

Daytime sleepiness increased during CPAP withdrawal and improved after CPAP re-initiation (ESS: 6.19 ± 3.11 at V1, 7.95 ± 5.02 at V2, and 5.60 ± 3.41 at V3, V1 vs V2 and V2 vs V3 0.001, Table 2). No serious adverse events related to the intervention were reported.

## Discussion

### Main findings

In this CPAP withdrawal and re-initiation study, two weeks off CPAP led to a marked recurrence of OSA with an impact observed on AHI, HB, PWAD index, and endothelial function (FMD) but only minor non-significant variations on autonomic markers such as BRS and on ΔHR. There was also a significant increase in diastolic blood pressure, in subjective sleepiness (ESS) and in some HRV indices along with reduction in autonomic response to cold pressor test.

### Endothelial and baroreflex outcomes

Changes in FMD reached statistical significance after CPAP re-initiation compared with the withdrawal visit suggesting some short-term effects of OSA recurrence on vascular function and some reversibility after two weeks of CPAP. Although a minimal clinically important difference has not been established for FMD, the 0.96% absolute increase observed after CPAP re-initiation is within the range associated with meaningful differences in the risk of incident CV events in prospective meta-analyses (8 to 13% decrease in risk per 1% higher FMD)[41–43]. Interestingly, the participants mean FMD (3.23 ± 2.49%) at V2 is below the 10^th^ percentile of a healthy population [44].

These findings are consistent with prior work using a CPAP withdrawal model to reveal short-term, reversible physiological consequences of untreated OSA. In the randomised CPAP withdrawal trial by Kohler and colleagues, withdrawal rapidly reinstated OSA and was accompanied by a significant deterioration in endothelial function assessed by FMD over a two-week period [28]. Our results are however relatively modest compared with this study. Several factors may contribute: OSAVE participants had lower baseline FMD, suggesting limited endothelial reserve that may limit detectability of further short-term deterioration. Differences in baseline treatment exposure, participant risk profiles, timing of assessment, and measurement variability may all contribute to between-study heterogeneity in the magnitude of FMD change.

Evidence from longer treatment horizons in minimally symptomatic OSA also support the concept of an impact of OSA on endothelial function. In a subset of the MOSAIC study with FMD measurements, six months of CPAP was associated with improvement in endothelial function [27], although phenotype heterogeneity remained important considerations. Overall, systematic reviews suggest that CPAP can improve endothelial function, while highlighting substantial variability across studies and populations, as well as sensitivity to acquisition and analytic protocols [45].

For BRS, the absence of statistically significant changes may reflect limited power and the long-standing CPAP exposure at baseline. In the literature, CPAP-naive patients with severe OSA, sustained CPAP has been reported to improve baroreflex control, supporting reversibility with longer treatment exposure [23, 29]. A plausible interpretation of the non-significant variation observed in this study is that participants entered the baseline visit in a comparatively recovered autonomic state after months/years of CPAP treatment and that two weeks without CPAP was insufficient to significantly impair baroreflex function or that this study sample size is too small to detect the actual effect size. Clinically, these results raise the hypothesis that some CPAP-associated autonomic improvements may be relatively durable, although larger studies may be required to detect more subtle changes.

### New OSA-related CV risk markers

Hypoxic burden, quantifying the integrated hypoxaemic exposure, showed a marked recurrence during CPAP withdrawal and improvement after treatment re-initiation, consistent with prior CPAP-withdrawal and treatment studies demonstrating that hypoxic burden is a treatment-responsive exposure metric that complements frequency-based indices such as the AHI [46, 47].

PWADi was also strongly treatment-state dependent, suggesting that peripheral vasoreactivity captured by PPG amplitude drops is rapidly modulated by OSA recurrence and treatment. The divergent behaviour of total PWADi and spontaneous PWADi is informative. The increase in total PWADi may be driven by respiratory-event related arousals and accompanying sympathetic surges, such that recurrence of apnoeas inflates event-associated vasoconstrictive episodes. However, this observation is discordant with recent cohort studies reporting that lower PWADi is associated with higher incident CV risk [18]. Therefore, the PWADi increase with OSA recurrence should not be interpreted as a sign of improvement of cardiovascular health.

On the other hand, spontaneous PWADi (uncoupled from respiratory events) decreased as AHI and HB increased off-CPAP. Consequently, the spontaneous PWAD events, which represents about 90% of all PWAD, may be a more informative marker of vasoreactivity capacity, that is not artificially inflated by the presence of respiratory perturbation.

Together with a trend towards lower BRS and the impairment in endothelial function (FMD), it raises the hypothesis that reductions in spontaneous PWADi may reflect concomitant deterioration in autonomic baroreflex control and endothelial-dependent vascular responsiveness during short-term untreated OSA.

Another possible explanation is that recurrence of OSA may increase the arousal threshold, as previously reported [48], which could reduce spontaneous arousals and consequently contribute to fewer spontaneous peripheral vasoconstrictive events captured by spontaneous PWADi.

These interpretations should be considered cautiously, as changes in spontaneous PWADi were small and not consistently significant across analyses.

ΔHR did not show a clear visit-to-visit effect, but missingness under CPAP limits inference. In exploratory within-person modelling, changes in ΔHR appeared more closely related to changes in hypoxic burden than changes in AHI (Table 3), suggesting that metrics capturing event severity rather than frequency alone may better reflect the physiological perturbation accompanying respiratory events, consistent with the hypothesis that intermittent hypoxaemia contributes to post-event heart rate acceleration [17, 47]. These results are hypothesis-generating and suggest also that robust evaluation of ΔHR in treated-state conditions may require methods tolerant to low event rates or alternative computation strategies.

Taken together, the differential responses to CPAP treatment across hypoxaemic exposure, vascular reactivity, and autonomic domains suggest that a multi-domain assessment may better capture OSA-related CV stress than any single severity metric.

### Secondary autonomic and vascular outcomes

#### HRV findings: apparent paradox and the potential value of non-linear indices

In the same way as total PWADi (see above), the increase in several conventional nocturnal HRV indices off CPAP is counterintuitive if interpreted as improved autonomic health. A more plausible explanation is that recurrent respiratory events, arousals, and state instability inflate conventional time- and frequency-domain HRV measures, limiting their validity as health surrogates in active OSA, as shown by Roche et al. by using these indices to screen for OSA [49]. The absence of parallel changes in daytime HRV supports this interpretation. In contrast, the relative stability of selected non-linear indices such as detrended fluctuation analysis (i.e. DFA α2) [50] suggests that these measures may be less sensitive to event-driven artefacts and could provide complementary information on autonomic regulation that is less dependent on OSA severity. This hypothesis requires validation with careful control of sleep stage distribution, artefact handling, and assessment of incremental prognostic value

#### Vascular structural parameters

Structural vascular parameters remained unchanged across visits, including measures of arterial stiffness and carotid structure (carotid elasticity and intima-media thickness). This null finding is biologically plausible given the short exposure windows in OSAVE. Structural vascular remodelling and carotid IMT reduction are generally reported over months rather than weeks on CPAP. Meta-analytic evidence suggests that carotid IMT reduction with CPAP is most apparent in studies with treatment durations of at least six months, with heterogeneous effects across populations [51]. meta-analytic evidence suggests that CPAP can improve arterial stiffness overall, but treatment effects are heterogeneous and are also typically evaluated over longer treatment durations than two weeks [52, 53].

#### Cold pressor test response

The systolic and diastolic blood pressure response to CPT was significantly lower after CPAP withdrawal suggesting a shift towards blunted autonomic and vascular reactivity during short-term untreated OSA. This pattern is consistent with evidence that OSA is associated with altered physiological and neural responses to autonomic challenges, including cold pressor stimulation [54, 55] . The incomplete recovery of cold pressor responses after 2 weeks of CPAP re-initiation aligns with evidence that autonomic alterations in OSA can persist despite treatment and may normalise gradually and incompletely, even over months, depending on phenotype and treatment exposure [23, 29].

#### Diastolic change in blood pressure

We observed a significant increase diastolic BP after CPAP withdrawal compared to V1 and V3, whereas systolic changes were more limited. These results are compatible with randomised CPAP withdrawal data showing that short-term withdrawal leads to measurable increases in morning blood pressure, and that office measurements may underestimate these effects compared with home or ambulatory monitoring [30, 56, 57]. Moreover, a recent post hoc analysis of a double-blind two-week CPAP withdrawal randomised controlled trial also reported that CPAP reduces morning diastolic BP and that this reduction was better predicted by AHI than by HB [46].

### Clinical implications

In this CPAP withdrawal and re-initiation model, key OSA-related CV risk markers were clearly treatment-responsive, but their trajectories were not uniform. Hypoxic burden showed a marked recurrence and reversal, total PWADi increased off CPAP and normalised after re-initiation, whereas spontaneous PWADi tended to change in the opposite direction. This divergence reinforces that these metrics should not be interpreted as interchangeable surrogates of OSA severity. Rather, they likely capture complementary dimensions of OSA pathophysiology, integrating exposure intensity (for example hypoxic burden) with individual vulnerability expressed through vascular and autonomic phenotypes, potentially reflected by PWAD-derived indices or ΔHR.

This study also showed short-term benefits of CPAP on endothelial function (FMD), diastolic blood pressure, autonomic reactivity to the CPT, and a clinically significant improvement in subjective sleepiness [58], which may also be associated with a reduction in long-term CV complications [59]. However, the inconsistent changes in other mechanistic vascular and autonomic endpoints suggest that a two-week exposure window, even with excellent adherence to CPAP, may be insufficient to capture slower adaptive vascular or autonomic processes in a previously treated, high-risk cohort.

Taken together, the heterogeneous responsiveness across OSA-related CV risk markers, and across autonomic and vascular results, supports an approach that integrates exposure metrics with markers of physiological response, rather than relying on a single severity metric, particularly in minimally symptomatic OSA where treatment targeting remains challenging.

Finally, although HRV is increasingly used as a surrogate of CV autonomic function, its interpretation in untreated OSA requires caution. The paradoxical rise in several conventional nocturnal HRV indices off CPAP likely reflects respiratory events, arousals, and sleep instability rather than improved autonomic health, especially given the lack of corresponding daytime changes. The relative stability of selected non-linear indices further suggests that HRV metric choice and clinical context are critical when using HRV to characterise autonomic function in OSA.

### Strengths, limitations, and future directions

Strengths of OSAVE include the standardised experimental protocol including within-subject withdrawal and re-initiation design, the clear recurrence and reversal of hypoxaemia and hypoxic burden confirming intervention fidelity with very high adherence to CPAP during the intervention and the integration of the main overnight OSA-related marker with next-morning broad vascular and autonomic phenotyping.

Several limitations should be emphasised. The design was non-randomised and open-label, and the exposure window was short, because prolonged CPAP withdrawal was considered ethically unacceptable. The study assessed multiple exploratory endpoints, and although domain-wise false discovery rate control was applied for secondary outcomes, residual multiplicity concerns remain. Endpoint-specific missingness was substantial for some measures, particularly ΔHR under CPAP and imaging-derived vascular outcomes such as FMD, despite adherence to contemporary measurement guidelines [26].

Finally, generalisability may be limited by the cohort’s high cardiometabolic risk burden and long-standing CPAP exposure, which may attenuate short-term physiological changes relative to CPAP-naive populations. The sex imbalance also limits the generalizability of the results.

Future investigations should determine whether autonomic and vascular phenotypes, together with emerging OSA-related CV risk markers, exhibit meaningful trajectories over longer periods of untreated exposure and following treatment initiation, and whether these measures provide incremental prognostic information beyond AHI. Larger studies will be needed to develop and validate integrated approaches that combine complementary markers such as exposure metrics extending beyond event frequency (for example hypoxic burden) with metrics that may capture host susceptibility (such as ΔHR and PWAD-derived indices), thereby capturing inter-individual heterogeneity in autonomic and vascular phenotypes.

## Conclusion

Even short-term re-exposure to untreated OSA was associated with early, modest but significant variations in FMD and diastolic BP suggesting vascular functional alterations over short time frames, with rapid reversibility. OSAVE also revealed a temporal dissociation between exposure and cardiovascular adaptation: AHI and HB responded rapidly to CPAP, whereas vascular structural and autonomic markers such as BRS showed only limited or no short-term change. These findings also highlighted the need for cautious interpretation of OSA-related markers during changes in AHI, as some indices such as total PWADi or HRV may misleadingly suggest cardiovascular improvement while primarily reflecting event-related variability. Markers less directly driven by respiratory events, such as spontaneous PWADi, may better capture underlying vascular or autonomic vulnerability.

## Supporting information

CONSORT Checklist

## Data Availability

De-identified individual participant data underlying the results reported in this article (including the data dictionary) will be made available upon reasonable request, subject to approval by the relevant ethics/governance bodies and the execution of a data sharing agreement.

## Acknowledgements

We thank Dr Hakim Khettab, Prof Rosa Maria Bruno and Prof Pierre Boutouyrie (Hôpital Européen Georges-Pompidou, AP-HP, Paris, France) for their support and for teaching the flow-mediated dilation technique, and Gianpaolo Lecciso (Centre d’Investigation et de Recherche sur le Sommeil, Lausanne University Hospital, Switzerland) for his contribution to the sleep research procedures. We also acknowledge the clinical research support from the Unité de Recherche Clinique from Anaesthesiology Unit - Lausanne University Hospital and especially Mrs Carine Marcucci. Artificial intelligence-assisted tools were used to support editing for grammar, spelling, and formatting. The authors retain full responsibility for the scientific content, data interpretation, and conclusions of this manuscript.

