## Supplementary material for "Effects of CPAP on OSA-related cardiovascular risk markers: a two-week CPAP withdrawal and re-initiation study": Online supplement

1. **Supplementary Methods**
   a. **Nonin WristOx2 3150 specifications** (Nonin, Plymouth, USA)

According to manufacturer specifications, the WristOx2 3150 reports SpO₂ and pulse rate using an exponential moving average with an effective window of approximately 4 beats. SpO₂ and pulse rate were exported with a sampling frequency of 3 Hz, and the photoplethysmography (PPG) pulse waveform was exported at 75 Hz. The wavelength used for pulse waveform is the infrared signal (910nm). Filtering is performed at acquisition and post-processing using proprietary, undisclosed manufacturer algorithms.

b. **Flow-mediated dilation**

Participants were studied supine in a quiet room after >15 min of rest, with the arm positioned at heart level. Baseline brachial diameter was recorded for 1 min prior to cuff inflation. A distal forearm cuff was inflated to 50 mmHg above systolic BP for 5 min and then rapidly deflated. Arterial diameter was continuously tracked for 4 min post-deflation to determine peak diameter and time-to-peak. Continuous diameter tracking and automated peak detection were performed using edge-detection software (FMD Studio©, Quipu, Pisa, Italy) with a stereotactic probe holder to reduce operator dependence. Mean blood velocity was obtained from the Doppler signal (with insonation angle kept constant at 72°) and shear rate (SR) was computed automatically as SR = 4 × Vmean / D, where Vmean is mean blood velocity and D is instantaneous arterial diameter. The hyperaemic stimulus was summarised as shear rate area-under-the-curve (AUC) from cuff release to the time of peak diameter. FMD was expressed as relative (%) change from baseline diameter. Endothelium-independent dilation was assessed once at V1 after sublingual nitroglycerin (400 µg), when not contraindicated, to quantify vascular smooth muscle responsiveness. All FMD recordings underwent quality control before analysis. FMD and associated variables (baseline diameter, peak diameter, time-to-peak and shear-rate AUC) were coded as missing when image quality was insufficient to ensure reliable edge detection and tracking (e.g., major motion artefacts, poor vessel delineation, or sustained loss of automated wall tracking), the proportion of missing FMD outcomes due to image quality is reported in Table S1.

c. **Bilateral carotid stiffness**

Bilateral carotid stiffness was assessed by ultrasound (Philips InnoSight, linear probe L12-4) using Carotid Studio© (Quipu, Pisa, Italy), enabling continuous tracking of common carotid artery (CCA) diameter and intima-media thickness (IMT). Measurements were performed in the supine position at a standardised segment of the common carotid artery (approximately 1-2 cm proximal to the bifurcation) on both sides. Estimated central systolic and diastolic blood pressures obtained immediately before the carotid ultrasound using the pOpmètre® system were used for pressure-calibrated stiffness indices. Carotid Studio© provided IMT (mm) and automatically derived stiffness metrics including local pulse wave velocity (m/s), compliance, distensibility and Young’s modulus. All carotid recordings underwent quality control before analysis. Measurements were excluded when image quality was insufficient to ensure reliable intima-media detection and tracking (e.g., major movement artefacts, poor vessel delineation, or sustained loss of automated wall tracking). The proportion of missing carotid outcomes due to image quality is reported in Table S1.

d. **Baroreflex sensitivity (BRS)**

After visual inspection and artefact correction the beat-by-beat values of systolic blood pressure (SBP) derived from the continuous BP recordings, and normal-to-normal intervals (NNI) from the ECG leads, were interpolated evenly at 5 Hz for spectral analysis. The sensitivity of the baroreflex control of heart rate was estimated by the transfer function technique (HLF). As to the SBP and NNI power spectra, and SBP-NNI cross-spectrum and coherency spectrum were calculated from the evenly resampled series at 5 Hz using 50% overlapped Hann data windows of 120 s length. The ratio between the SBP-NNI cross-spectrum and NNI spectrum was calculated considering again only spectral lines with squared coherence modulus > 0.2 and averaged over the LF band, providing the HLF estimate of BRS.

2. **Supplementary Tables**

**Table S1. Missing data by visit**

| **Group** | **Outcome** | **Missing V1 (n)** | **Missing V2 (n)** | **Missing V3 (n)** |
| --- | --- | --- | --- | --- |
| **Polygraphy** | AHI^†^ (events/h) | 1 | 0 | 0 |
|  | ODI^†^ (desat/h) | 1 | 0 | 0 |
|  | Mean SpO2^†^ (%) | 3 | 0 | 0 |
|  | T90 (min) | 1 | 0 | 0 |
|  | Mean desaturation (%) | 3 | 0 | 0 |
|  | Estimated TST (min) | 1 | 0 | 0 |
| **New OSA-related CV metrics** | HB^†^ (%·min/h) | 3 | 0 | 0 |
|  | ΔHR^†^ (beats/min) | 29 | 1 | 28 |
|  | ΔHR (std)^†^ | 29 | 1 | 28 |
|  | PWADi^†^ (drops/h) | 3 | 0 | 0 |
|  | PWADi spontaneous^†^ (drops/h) | 3 | 0 | 0 |
|  | Spontaneous PWADi / duration (drops·h⁻¹·s⁻¹) | 5 | 0 | 5 |
|  | Mean drops duration (s) | 3 | 0 | 0 |
|  | Mean drops descending slope (%/s) | 6 | 2 | 2 |
|  | Mean drops ascending slope (%/s) | 5 | 1 | 1 |
| **Vascular parameters** | FMD^†^ (%) | 12 | 8 | 5 |
|  | FMD recovery (%) | 8 | 4 | 3 |
|  | RCCA mean IMT (mm) | 7 | 8 | 3 |
|  | LCCA mean IMT (mm) | 12 | 11 | 7 |
|  | RCCA stiffness (m/s) | 7 | 8 | 3 |
|  | LCCA stiffness (m/s) | 12 | 11 | 7 |
|  | RCCA Young modulus (kPa) | 7 | 8 | 3 |
|  | LCCA Young modulus (kPa) | 12 | 11 | 7 |
|  | Peripheral Pulse Wave Velocity (m/s) | 2 | 2 | 3 |
| **Diurnal autonomic parameters** | Supine BRS^†^ (ms/mmHg) | 3 | 4 | 6 |
|  | Seated BRS (ms/mmHg) | 4 | 3 | 4 |
|  | Supine BEI | 9 | 5 | 11 |
|  | Seated BEI | 5 | 4 | 4 |
|  | Supine Latency + (s) | 17 | 15 | 21 |
|  | Seated Latency + (s) | 17 | 11 | 14 |
|  | Supine Latency - (s) | 14 | 16 | 20 |
|  | Seated Latency - (s) | 13 | 14 | 10 |
|  | Diurnal HRV - VLF (ms^2^) | 0 | 1 | 0 |
|  | Diurnal HRV - LF (ms^2^) | 0 | 1 | 0 |
|  | Diurnal HRV - HF (ms^2^) | 0 | 1 | 0 |
|  | Diurnal HRV - LF/HF | 0 | 1 | 0 |
|  | Reaction to CPT - HR (bpm) | 2 | 2 | 2 |
|  | Reaction to CPT - SBP (mmHg) | 2 | 2 | 2 |
|  | Reaction to CPT - DBP (mmHg) | 2 | 2 | 2 |
| **Nocturnal autonomic parameters** | SDNN (ms) | 1 | 0 | 0 |
|  | RMSSD (ms) | 1 | 0 | 0 |
|  | VLF (ms^2^) | 1 | 1 | 0 |
|  | LF (ms^2^) | 1 | 1 | 0 |
|  | HF (ms^2^) | 1 | 1 | 0 |
|  | LF/HF | 1 | 1 | 0 |
|  | Acceleration Capacity (ms) | 1 | 1 | 0 |
|  | Deceleration Capacity (ms) | 1 | 1 | 0 |
|  | DFA α1 | 1 | 1 | 0 |
|  | DFA α2 | 1 | 1 | 0 |
|  | SampEn | 20 | 19 | 20 |
| **Blood pressures** | SBP - armcuff - seated (mmHg) | 0 | 0 | 0 |
|  | DBP - armcuff - seated (mmHg) | 0 | 0 | 0 |
|  | SBP - digit finapres - supine 10 min (mmHg) | 0 | 0 | 0 |
|  | DBP - digit finapres - supine 10 min (mmHg) | 0 | 0 | 0 |
|  | SBP - digit finapres - seated 10 min (mmHg) | 0 | 0 | 0 |
|  | DBP - digit finapres - seated 10 min (mmHg) | 0 | 0 | 0 |
| **Weight** | BMI (kg/m^2^) | 0 | 0 | 0 |
| **Sleepiness** | Epworth Score | 0 | 0 | 0 |

**Table S1: Missing data by visit -** Number of missing observations for each endpoint at each visit. Missing data were not imputed.

**Abbreviations:** V1, visit 1; V2, visit 2; V3, visit 3; CPAP, continuous positive airway pressure; AHI, apnoea-hypopnoea index; ODI, oxygen desaturation index; SpO₂, peripheral oxygen saturation; T90, time spent with SpO₂ <90%; TST, total sleep time; CV, cardiovascular; HB, hypoxic burden; ΔHR, event-related heart rate response; ΔHR (std), event-related heart rate response adjusted for baseline heart rate; PWADi, pulse wave amplitude drop index; FMD, flow-mediated dilation; RCCA, right common carotid artery; LCCA, left common carotid artery; IMT, intima-media thickness; BRS, baroreflex sensitivity; BEI, baroreflex effectiveness index; HRV, heart rate variability; VLF, very low frequency; LF, low frequency; HF, high frequency; CPT, cold pressor test; HR, heart rate; SBP, systolic blood pressure; DBP, diastolic blood pressure; SDNN, standard deviation of normal-to-normal intervals; RMSSD, root mean square of successive differences; DFA, detrended fluctuation analysis; DFA α1, short-term scaling exponent; DFA α2, long-term scaling exponent; SampEn, sample entropy; BMI, body mass index

**Table S2. Baroreflex latency and continuous blood pressure across study visits**

| **Group** | **Outcome** | **Visit 1 - ON CPAP Value** | **Visit 2 - OFF CPAP Value** | **Visit 3 - ON CPAP Value** | **p-value V1 vs V2** | **p-value V2 vs V3** | **p-value V1 vs V3** |
| --- | --- | --- | --- | --- | --- | --- | --- |
| **Diurnal autonomic parameters** | Supine Latency + (s) | 0.80 (0.37) | 0.77 (0.39) | 0.87 (0.58) | 0.886 | 0.905 | 0.998 |
|  | Seated Latency + (s) | 0.79 (0.40) | 0.86 (0.41) | 0.71 (0.32) | 0.886 | 0.611 | 0.998 |
|  | Supine Latency - (s) | 0.89 [0.64, 1.02] | 0.77 [0.60, 1.10] | 0.86 [0.66, 1.11] | 0.886 | 0.937 | 0.998 |
|  | Seated Latency - (s) | 0.80 [0.67, 1.00] | 0.83 [0.54, 1.02] | 0.87 [0.58, 1.16] | 0.886 | 0.611 | 0.998 |
| **Blood pressures** | SBP - digit finapres - supine 10 min (mmHg) | 140.25 (13.97) | 138.96 (15.51) | 135.62 (16.56) | 0.698 | 0.101 | 0.143 |
|  | DBP - digit finapres - supine 10 min (mmHg) | 82.14 [75.32, 85.97] | 81.71 [77.64, 91.88] | 78.88 [72.64, 84.93] | 0.256 | **0.006**** | 0.262 |
|  | SBP - digit finapres - seated 10 min (mmHg) | 138.38 [130.71, 151.39] | 138.88 [129.08, 150.36] | 138.43 [129.85, 147.44] | 0.762 | 0.101 | 0.262 |
|  | DBP - digit finapres - seated 10 min (mmHg) | 77.41 [72.74, 85.92] | 79.76 [71.90, 86.11] | 76.19 [72.39, 83.64] | 0.673 | **0.044*** | 0.262 |

**Table S2: secondary endpoints across study visits –** P-values are Benjamini–Hochberg FDR-adjusted within each outcome domain (Group). SBP and DBP correspond to the mean blood pressure derived from continuous finger arterial pressure recordings (Finapres) over a 10-min period. Data are reported as mean (SD) or median [IQR], as appropriate. Pairwise between-visit differences were estimated using linear mixed-effects models adjusted for age, sex, and BMI, with a participant-specific random intercept; p-values are from model-based contrasts. Significance: *p<0.05, **p<0.01, ***p<0.001

**Abbreviations:** V1, visit 1; V2, visit 2; V3, visit 3; CPAP, continuous positive airway pressure; SBP, systolic blood pressure; DBP, diastolic blood pressure

**Table S3. Within-patient associations of baroreflex latency and continuous blood pressure with changes in AHI and hypoxic burden.**

| **Group** | **Outcome** | **AHI within β per 10 units [95% CI]** | **p-value** | **HB within β per 10 units [95% CI]** | **p-value** |
| --- | --- | --- | --- | --- | --- |
| Diurnal autonomic parameters | Supine Latency + (s) | -0.010 [-0.055, 0.036] | 0.725 | -0.000 [-0.021, 0.021] | 0.979 |
|  | Seated Latency + (s) | 0.038 [-0.005, 0.081] | 0.327 | 0.013 [-0.003, 0.029] | 0.711 |
|  | Supine Latency - (s) | 0.040 [-0.016, 0.096] | 0.445 | 0.021 [-0.002, 0.044] | 0.711 |
|  | Seated Latency - (s) | -0.043 [-0.089, 0.003] | 0.327 | -0.003 [-0.020, 0.014] | 0.979 |
| Blood pressures | SBP - digit finapres - supine 10 min (mmHg) | 0.360 [-0.675, 1.395] | 0.491 | 0.071 [-0.331, 0.472] | 0.790 |
|  | DBP - digit finapres - supine 10 min (mmHg) | 0.858 [0.176, 1.540] | **0.043*** | 0.265 [-0.001, 0.532] | 0.153 |
|  | SBP - digit finapres - seated 10 min (mmHg) | 0.980 [-0.095, 2.055] | 0.147 | 0.151 [-0.274, 0.575] | 0.723 |
|  | DBP - digit finapres - seated 10 min (mmHg) | 0.552 [-0.201, 1.305] | 0.222 | 0.148 [-0.144, 0.440] | 0.634 |

**Table S3: Within-patient associations of secondary endpoints with changes in AHI and hypoxic burden -** Associations were estimated using mixed models adjusted for age, sex and BMI, with a participant-specific random intercept, including a within–between decomposition of AHI and HB: the within-person component represents deviation from each participant’s mean across visits, and the between-person component represents participant-specific means. β coefficients are shown per 10-unit increase in within-person AHI (events/h) or within-person HB (%·min/h)**.** P-values are Benjamini–Hochberg FDR-adjusted within each outcome domain (Group). P-values Significance: * p<0.05, ** p<0.01, *** p<0.001.

**Abbreviations:** AHI, apnoea-hypopnoea index; HB, hypoxic burden; SBP, systolic blood pressure; DBP, diastolic blood pressure
